# Identification of Multi-repeat Sequences using Genome Mining Approaches for Developing Highly Sensitive Molecular Diagnostic Assay for the Detection of *Chlamydia trachomatis*

**DOI:** 10.1101/2023.07.19.22272924

**Authors:** Clement Shiluli, Shwetha Kamath, Bernard N. Kanoi, Rachael Kimani, Michael Maina, Harrison Waweru, Moses Kamita, Ibrahim Ndirangu, Hussein M. Abkallo, Bernard Oduor, Nicole Pamme, Joshua Dupaty, Catherine M. Klapperich, Srinivasa Raju Lolabattu, Jesse Gitaka

**Author notes:** **Corresponding authors:** Jesse Gitaka, Srinivasa Raju Lolabattu.

## Abstract

*Chlamydia trachomatis (C. trachomatis)* is a common sexually transmitted infection (STI). In 2019, the World Health Organization reported about 131 million infections. The majority of infected patients are asymptomatic with cases remaining undetected. It is likely that missed *C. trachomatis* infections contribute to preventable adverse health outcomes in women and children. Consequently, there is an urgent need of developing efficient diagnostic methods. In this study, genome-mining approaches to identify identical multi-repeat sequences (IMRS) distributed throughout the *C. trachomatis* genome were used to design a primer pair that would target regions in the genome. Genomic DNA was 10-fold serially diluted (100pg/μL to 1×10-3pg/μL) and used as DNA template for PCR reactions. The gold standard PCR using 16S rRNA primers was also run as a comparative test, and products were resolved on agarose gel. The novel assay, *C. trachomatis* IMRS-PCR, had an analytical sensitivity of 9.5 fg/μL, representing better sensitivity compared with 16S rRNA PCR (4.31 pg/μL). Our experimental data demonstrate the successful development of lateral flow and isothermal assays for detecting *C. trachomatis* DNA with potential use in field settings. There is a potential to implement this concept in miniaturized, isothermal, microfluidic platforms, and laboratory-on-a-chip diagnostic devices for reliable point-of-care testing.

## 2. Introduction

*Chlamydia trachomatis* (*C. trachomatis*) is one of the most common sexually transmitted infections (STIs) associated with adverse birth and neonatal outcomes such as preterm labor with low birth weight (1). During delivery, the risk of vertical transmission of *C. trachomatis* increases (2). Most patients with *C. trachomatis* infections are asymptomatic, and these cases remain undetected and untreated, further complicating the standard of care in most countries (3). It is, therefore, likely that missed maternal *C. trachomatis* infections contribute to preventable adverse health outcomes among women and children globally, such as endometritis, salpingitis, tubo-ovarian abscesses, pelvic peritonitis, and perihepatitis (4). In 2019 the World Health Organization (WHO) reported about 131 million cases of *C. trachomatis* globally (5).

Traditional methods of *C. trachomatis* testing include cervical cytological examination using Papanicolaou (Pap) smears (6). However, these techniques have many challenges, such as technical difficulties during cultures, labor intensiveness, increased turnaround time, and high cost. *C. trachomatis* infections are diagnosed by direct and indirect methods (7).

Direct methods detect the presence of *C. trachomatis* in localized infections (7). These methods include culture, antigen tests (Enzyme Immuno Assays (EIA), Direct Fluorescent Antibody (DFA) tests, immune chromatographic tests, Rapid Detection Tests (RDTs), and Nucleic Acid Amplification Tests (NAATs) (7).

Indirect methods are specific to *C. trachomatis* antibodies and may be used for diagnostic evaluation of chronic/invasive infection (Pelvic Inflammatory Disease, lymphogranuloma venereum) and post-infectious complications, like sexually acquired reactive arthritis (SARA) (7).

Direct detection methods, such as detecting *C. trachomatis* using NAATs, have several advantages. They are based on polymerase chain reaction (PCR) and use fluorescence-labeled probes to detect amplification products in real-time, thereby significantly reducing the turn-around time (8). Studies have also shown that *C. trachomatis* PCR using DNA extracted from conjunctival swabs can achieve a detection limit of up to 100 plasmid copies (9).

Other NAATs, such as ligase chain reaction (LCR) and transcription-mediated amplification (TMA), are also sensitive and specific in the detection of *C. trachomatis* in clinical samples (10). Results of these tests can be generated in a few hours when coupled with automated nucleic acid extraction techniques.

However, NAATs such as the Abbot Real-Time CT/NG requires stringent sample transport and storage conditions (11). For example, to achieve accurate and reliable results, samples from asymptomatic women must be stored between 2°C and 30°C and processed within 14 days after collection (11). Symptomatic women’s specimens must be thawed, frozen, and stored at the same temperature range (11). These conditions may be challenging, mainly in resource-limited countries where refrigeration facilities are unavailable, especially during specimen collection and transport (12).

When compared to *C. trachomatis* culture, the DFA has a sensitivity of between 95% - 100% (13). However, DFA involves labor-intensive microscopic examination of individual stained specimens, which can be time-consuming when dealing with many samples (13). Also, highly skilled and experienced personnel perform routine microscopic examinations (13). In contrast, EIAs are ideal for testing large numbers of samples and achieve sensitivities of 90% in comparison with culture, but may be less sensitive than DFA and may give false positive tests (13).

Thriving *C. trachomatis* culture depends on isolated vital organisms, and the detection rate in clinical samples ranges between 60%–80%. In addition, culture sensitivity may be affected by inadequate specimen collection, storage, and transport, toxic substances in clinical specimens, and overgrowth of cell cultures by commensal microorganisms (14). *C. trachomatis* culture has an extended turn-around time; it is labor intensive, and different laboratories have various standardization protocols (14).

The use of indirect methods for the detection of *C. trachomatis* is inaccurate in identifying acute infections of the lower genital and anal tract; this is because antibody responses become detectable only after weeks to months and are often less pronounced (14).

Enzyme immunoassays based on the detection of bacterial lipopolysaccharide may cross-react with other gram-negative bacteria leading to false-positive results. The chlamydial antibody response is either delayed or absent in some individuals, which makes most serological tests inadequate (15). In addition, most the *C. trachomatis* infections are asymptomatic and are diagnosed late, resulting in uninterrupted transmission (16). To manage asymptomatic *C. trachomatis* infections, there is an urgent need to develop efficient diagnostic methods with adequate sensitivity and specificity, which, when implemented as part of screening programs, can contribute to identifying new cases and controlling the transmission of *C. trachomatis* infections (17). In this study, we developed a novel method based on de novo genome mining strategy that identifies identical multi-repeat sequences (IMRS) in *C. trachomatis* genome for use as both PCR and isothermal amplification assays. The assay has a potential for field deployability due to inherent high sensitivity.

## 3. Materials and Methods

### 3.1 IMRS genome mining algorithm

The primers used in this study were designed based on Identical Multi-Repeat Sequence (IMRS) genome mining algorithm as previously described (18). The *C. trachomatis* genome (NC_000117.1) was used as the reference to identify identical, repetitive sequence substrings of <30 bases that can be used as forward or reverse primers. NIH’s Basic Local Alignment Search Tool (BLAST) was used to evaluate the primer pairs to confirm *C. trachomatis* specificity. The resulting primers were predicted to amplify several fragments of DNA of different sizes derived from different regions of the *C. trachomatis* genome.

### 3.2 C. trachomatis Genomic DNA Preparation

*C. trachomatis* DNA was obtained from the American Type Culture Collection (ATCC) (ATCC® VR-885D™, LOT No. 70013611) at a concentration of ≥ 1 × 105 copies/μL. The stock DNA was diluted to a concentration of 100 pg/μL (8.92 × 104 copies/μL) and thereafter serially diluted 100 fold and 10 fold in TE buffer (Thermo Fisher Scientific, Waltham, Massachusetts, USA) for the *C. trachomatis* IMRS and 16S rRNA PCR, respectively.

### 3.3 16S rRNA PCR

The 16S rRNA PCR assays was carried out in a reaction mixture containing dNTPs (Thermo Fisher Scientific, Waltham, Massachusetts, USA) (0.2mM), forward and reverse primers (0.01 mM each), Taq Hot-Start DNA polymerase (Thermo Fisher Scientific, Waltham, Massachusetts, USA) (1.25 U), genomic template DNA, 1 μL to a final PCR reaction volume of 25 μL. The cycling parameters were as follows: 95°C for 3 min; 40 cycles of: 95°C for 30 s, 56°C for 30 s, 72°C for 30 s; and 72°C for 5 min and a final hold of 4°C.

### 3.4 C. trachomatis IMRS PCR

The *C. trachomatis* IMRS assays were carried out in a reaction mixture containing dNTPs (Thermo Fisher Scientific, Massachusetts, USA) (0.2 mM), forward and reverse primers (0.01 mM each), Taq Hot-Start DNA polymerase (Thermo Fisher Scientific, Massachusetts, USA) (1.25 U), genomic template DNA, 1 μL to a final PCR reaction of 25 μL. The cycling parameters for *C. trachomatis*-IMRS assay was as follows: 95°C for 3 min; 40 cycles of: 95°C for 30 s, 50°C for 30 s, 72°C for 30 s; and 72°C for 5 min and a final hold of 4°C. All PCR products were resolved in 1% agarose gel visualized on a UV Gel illuminator system (Fison Instruments, Glasgow, United Kingdom) under ethidium bromide staining.

### 3.5 Isothermal IMRS Amplification Assay

The Isothermal (Iso) IMRS amplification were performed in a 25 μL reaction mixture consisted of *Bst* 2.0 polymerase (640 U/mL) (New England Biolabs, Massachusetts, USA), with 1× isothermal amplification buffer, 3.2 μM forward primer, and 1.6 μM reverse primer (Jigsaw Biosolutions, Bengaluru, India) combined with 10 mM dNTPs (Thermo Fisher Scientific, Massachusetts, USA), 0.4 M Betaine (Sigma-Aldrich, Missouri, USA), molecular-grade water and Ficoll (0.4 g/mL) (Sigma-Aldrich, Missouri, USA). Amplification was carried out at 56°C for 40 min. Amplified products were visualized by gel electrophoresis in 1% gel.

### 3.6 Lower limit of detection

To assess the lower limit of detection (LLOD) of the *C. trachomatis* IMRS PCR assay, genomic DNA was diluted 100-fold from 100 pg/μL (8.92 × 104 copies/μL) to 10-6 pg/μL (<1 copies/μL) and 10-fold from 100 pg/μL (8.92 × 104 copies/μL) to 10-2 pg/μL (< 1 copies/μL) for the gold standard 16S rRNA PCR. Thereafter, 5 replicates of each dilution were used for the assays. Amplification products were visualized on gel after electrophoresis. To determine the LLOD of the *C. trachomatis* 16S rRNA and IMRS PCR, probit analysis was performed using the ratio of successful reactions to the total number of reactions performed for each assay. Similarly, to assess the LLOD for the *C. trachomatis* Iso IMRS PCR assay, genomic DNA was diluted 10-fold from a starting concentration of 1.64 × 106 copies/μL. Thereafter, five replicates of each dilution were used to determine the LLOD by using the ratio of successful reactions to the total number of reactions performed for each assay.

### 3.7 C. trachomatis-Lateral Flow Assay

To generate a visual read-out signal of amplicons, the *C. trachomatis*-Lateral Flow Assay (*C. trachomatis*-LFA) was used. Briefly, 2.5 μL annealing buffer, dNTPs and NaCl (1.75 μL), MgSO4 (1.2 μL), NG 5’ biotinylated forward primer (Biotin 5’-“TGCTGCTGCTGATTACGAGCCGA”-3’), *C. trachomatis* Reverse primer, *C. trachomatis* 3’ FAM labelled probe (CCACCAATACTCTC/-FAM-3’), *C. trachomatis* Digotexin labelled probe, Ficoll 400 (6.25 μL), *C. trachomatis* internal control sequence DNA (2.5 μL), ISO Amp III enzyme mix 2.0 μL, template DNA 5μL and molecular grade water 1.45 μL in a final master mix volume of 25 μL was used. Incubation of the LFA strip (Milenia Biotec GmbH, Giessen, Germany) was performed at 65°C for 1 hour and 5 #L of reaction mixture was then added to the LFA strip, thereafter, 2 drops of running buffer was also added.

### 3.8 C. trachomatis IMRS and 16S rRNA PCR real-time PCR assay

The Quant Studio 5 Real-Time PCR System (with Quant Studio Design and Analysis Desktop Software v1.5.) was used as a reference method (19) for the *C. trachomatis* 16S rRNA PCR and the *C. trachomatis* IMRS-PCR and for determining the sensitivity of the *C. trachomatis* IMRS and 16S rRNA PCR primers for detecting *C. trachomatis* DNA. The genomic DNA was serially diluted 10-fold starting concentration of 104 genome copies/μL. The final real-time PCR master mix volume was 10 μL in triplicate and consisted of the following components: 5 μL SYBR Green qPCR Master Mix (Thermo fisher, Massachusetts, USA), 1 μL forward and reverse IMRS primer mix, 2.5 μL template genomic DNA and 1.5 μL molecular grade water. The amplification cycling conditions were 50°C for 2 min; 95°C for 10 min; 40 cycles of 95°C for 15 s and 60°C for 30 s.

### 3.9 Cloning and characterization of *C. trachomatis*-IMRS amplicons

Gene cloning was performed to confirm the sequences of amplicons obtained from the *C. trachomatis* IMRS PCR assay. *Chlamydia trachomatis* gDNA was amplified using Assembly_IMRS-F (TTCCGGATGGCTCGAGTTTTTCAGCAAGATTGCCTGCCT**<u>GCTGATTACGAGCCG</u>**

**<u>A</u>**) and Assembly IMRS-R (AGAATATTGTAGGAGATCTTCTAGAAAGATT**<u>GTAGGAGGAGCCTCTTAGAGA A</u>**) primers. The underlined bold sequences correspond to the IMRS primers for amplifying the *C. trachomatis* genome whereas the bold underline in the primers corresponds to sequences in the cloning vector. The resulting amplicons was resolved on 2% agarose to confirm the fragment size and subsequently purified using the PureLink™ PCR purification kit (ThermoFisher). The purified amplicon was then ligated into pJET1.2/ blunt vector (ThermoFisher) using the NEBuilder® HiFi DNA Assembly kit (NEB) as per the manufacturer’s instruction. The resulting NEBuilder HiFi DNA Assembly product was transformed into NEB 5-alpha Competent *E. coli* (NEB #C2987, NEB) following the manufacturer’s instructions. Transformed colonies were randomly selected, DNA extracted and Sanger-sequenced using the universal pJET1.2 forward sequencing primer (CGACTCACTATAGGGAGAGCGGC) and pJET1.2 forward sequencing primer (AAGAACATCGATTTTCCATGGCAG). The resulting nucleotides were trimmed and analysed using SnapGene software (GSL Biotech; available at snapgene.com), aligned to check for similarity or “clonal” differences and BLAST used to check for similarity with the *C. trachomatis* genome.

### 3.10 Clinical samples

Vaginal swab samples collected from a cohort of women aged between 19 – 49 years enrolled in an STI screening study at the Kenyatta National Hospital in Nairobi County in 2022 were used in this study. The *C. trachomatis*-16S rRNA conventional PCR assay was used to confirm *C. trachomatis* infections. Thereafter, *C. trachomatis* positive samples were used to validate the *C. trachomatis*-IMRS primers. Participants were notified of results directly and confidentially by study staff, and were treated for *C. trachomatis* infection. Use of these samples and study proposal was approved by the Mount Kenya University Ethical Review Committee (MKU/ERC/1649).

### 3.11 Data analysis

Graphs were plotted with GraphPad Prism version 7.0 (GraphPad Software, San Diego, CA). The mean, and SD values were calculated with Excel 2016. To determine the LLOD of *C. trachomatis*-IMRS and *C. trachomatis*-16S rRNA PCR assays (the concentration at which genomic *Chlamydia trachomatis* DNA is detected with 95% confidence), probit regression analyses were performed in Excel 2016. Genomic *Trichomonas vaginalis* and *Treponema pallidum* DNA were used to determine the specificity of the *C. trachomatis*-IMRS primers for other STIs. Statistical analyses were performed using *t*-test of GraphPad Prism version 7.0. for two-tailed distribution. *P* < 0.05 was considered as significant.

## 4. Results

### 4.1 Design and Distribution of IMRS Primer Sets on *Chlamydia trachomatis* Genome

A total of 6 repeats (Table 1) were identified using the IMRS-based genome mining algorithm [10], which could be used as forward and reverse primers for an amplification assay. These selected primers (F 5’-TGCTGCTGCTGATTACGAGCCGA -3’ and R 5’-TGTAGGAGGAGCCTCTTAGAGAA - 3’), as were depicted using a circos plot (Fig 1), were found to be present at various loci of the sense and antisense strands, allowing them to serve interchangeably as forward or reverse primers.

**Table 1:** IMRS Primer target regions on the *Chlamydia trachomatis* bacterial genome

| No. | Amplification regions | Expected sequence fragment |
| --- | --- | --- |
| 1 | 531375 - 531511 | 137bp |
| 2 | 531375 - 531661 | 287bp |
| 3 | 531375 - 531814 | 440bp |
| 4 | 531525 - 531661 | 137bp |
| 5 | 531525 - 531814 | 290bp |
| 6 | 531675 - 531814 | 140bp |

**Fig 1:**
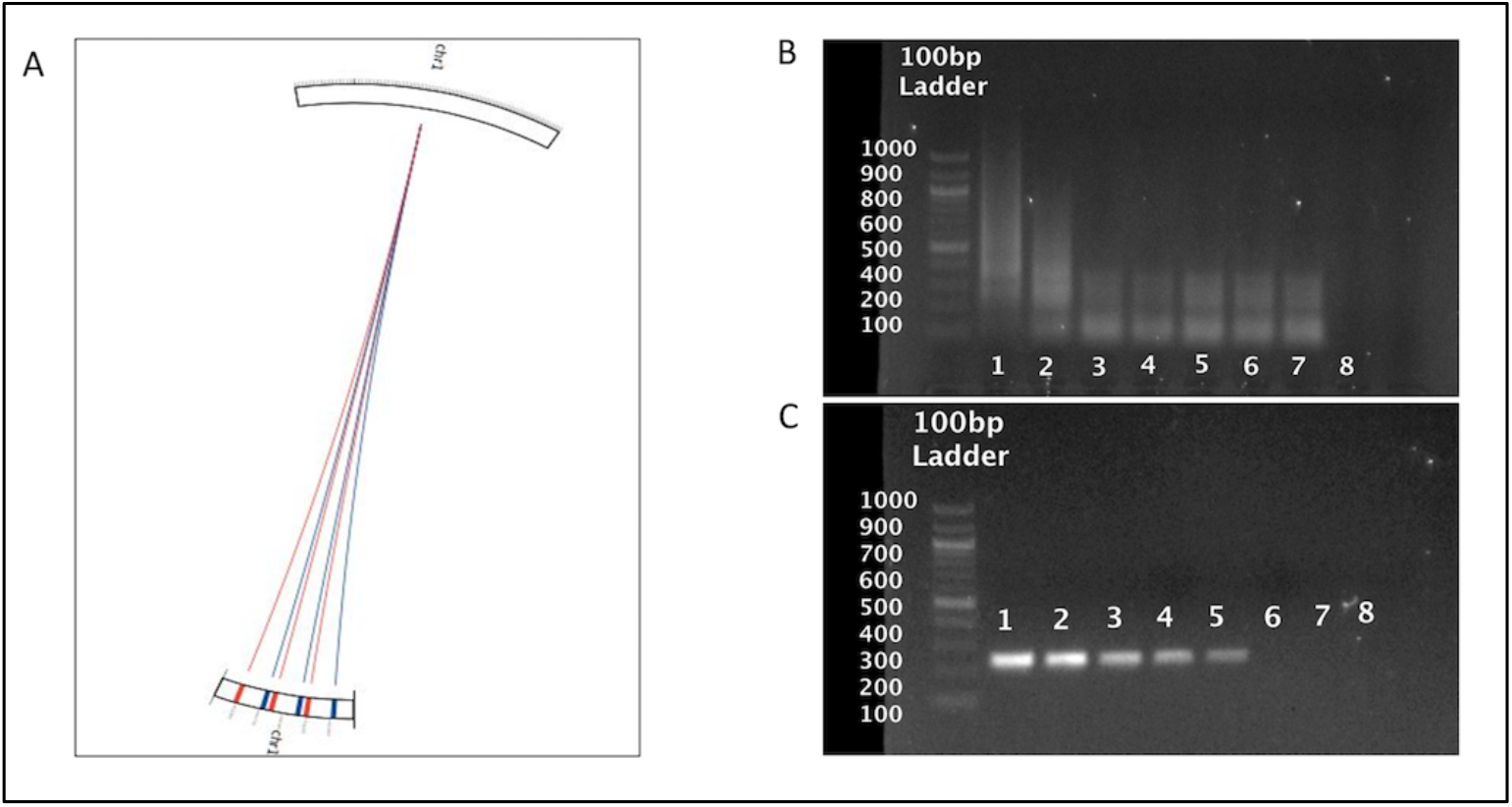
*C. trachomatis*-IMRS primer targets and gel images from the *C. trachomatis* IMRS and conventional 16S rRNA PCR assay**: (A)** Circos plot for the distribution of identical multi repeat sequence (IMRS) primers in the Chlamydia trachomatis genome. Chlamydia trachomatis IMRS primer A (blue lines) IMRS primer B (red lines) both have 6 repeats. Image of 10-fold serially diluted (1 – 100, 2 – 10, 3 – 1, 4 – 0.1, 5 – 0.01, 6 – 0.01, 7 – 0.001 and 8 - NTC) (pg/μl) genomic *Chlamydia trachomatis* DNA amplicons resolved on 1% gel using IMRS primers (B) and gold standard 16S rRNA PCR (C).

### 4.2 Amplification of Sequences on *Chlamydia trachomatis* Genome using Identical Multi-Repeat Primers and Gold Standard 16S rRNA PCR

To confirm the ability and specificity of the IMRS primers to amplify the targeted regions on the *C. trachomatis* genome, serially diluted DNA was used as a template for PCR amplification. The IMRS primers could detect *C. trachomatis* genomic DNA from a concentration of as low as 0.1 fg/μL (Fig 1B), whereas the gold standard 16S rRNA primers (Fig 1C) could only detect *C. trachomatis* genomic DNA down to a concentration of 10 fg/μL. This demonstrates that the IMRS-based PCR assay has a 100-fold higher sensitivity compared to the gold standard 16S rRNA PCR assay.

Real-time PCR was also performed using serially diluted genomic *C. trachomatis* DNA as a template and *C. trachomatis*-IMRS primers (Table 2A), as well as conventional *C. trachomatis*-16S rRNA primers (Table 2B). The mean Ct values at each respective dilution were used to plot amplification curves (Fig 2A for *C. trachomatis*-IMRS primers and Fig 2B for *C. trachomatis*-16S rRNA primers).

**Table 2:** Genomic DNA dilution to determine the sensitivity of the *C. trachomatis*-IMRS primers using Real time PCR. Serially diluted *Chlamydia trachomatis* genomic DNA served as amplification templates for the *C. trachomatis*-IMRS (A) and 16S rRNA primers (B).

**A**
| Concn of DNA (genome copies/μl) | Ct value | STD.DEV |
| --- | --- | --- |
| $1 \times 10^4$ | 12.298 | 0.541 |
| $1 \times 10^3$ | 17.674 | 1.244 |
| $1 \times 10^2$ | 23.639 | 0.349 |
| $1 \times 10^1$ | 28.265 | 0.275 |

**B**
| Concn of DNA (genome copies/μl) | Ct value | STD.DEV |
| --- | --- | --- |
| $1 \times 10^4$ | 14.64 | 0.902 |
| $1 \times 10^3$ | 20.584 | 1.599 |
| $1 \times 10^2$ | 26.607 | 1.056 |
| $1 \times 10^1$ | 36.527 | 0.249 |

**Fig 2:**
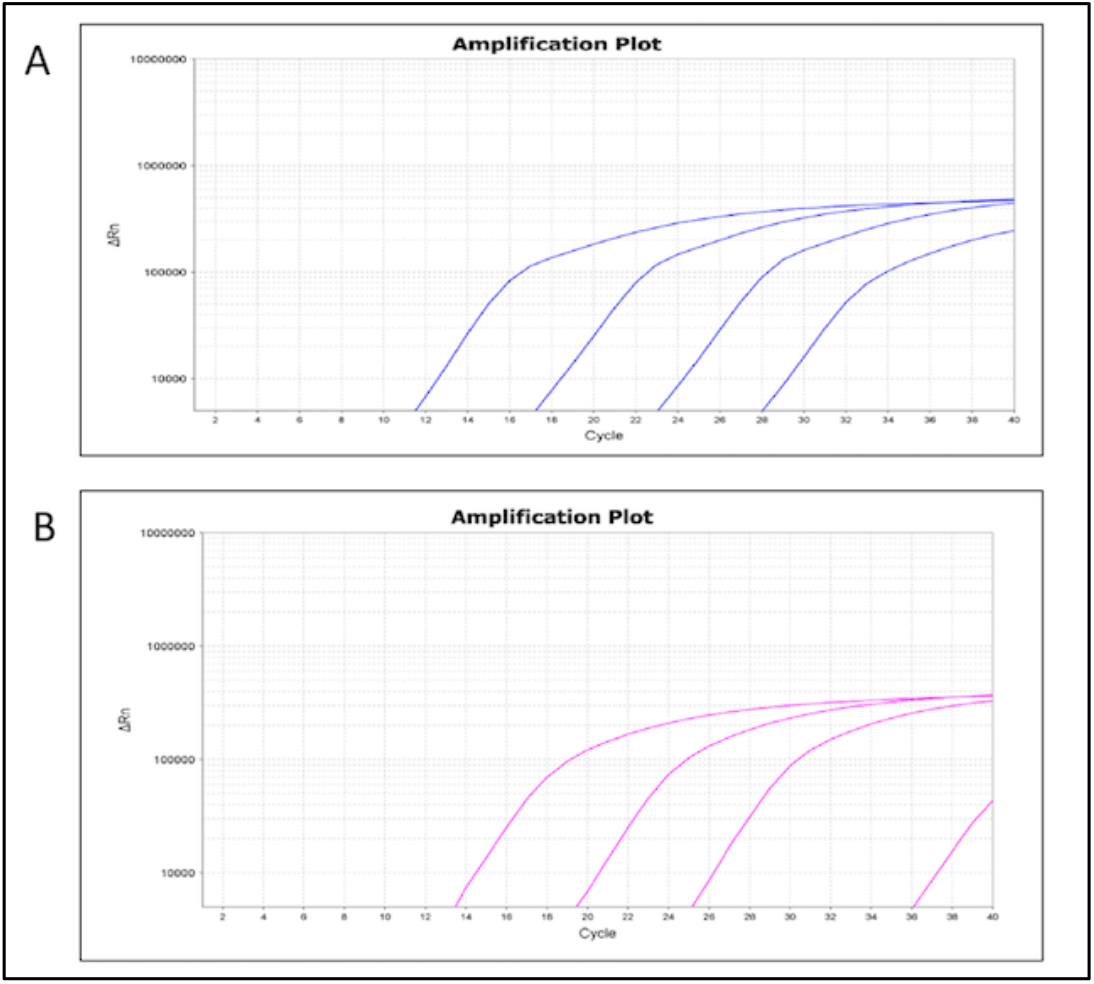
Real-time PCR amplification plots from the *C. trachomatis*- IMRS (A) and the 16S rRNA (B) assays using serially diluted *Chlamydia trachomatis* genomic DNA.

### 4.3 Determination of the Lower Limit of Detection (LLOD) for IMRS PCR and Gold Standard 16S rRNA PCR assay

To determine the lowest limit of detection (LLOD) of the IMRS PCR relative to the gold standard 16S rRNA PCR, probit statistic was performed using *C. trachomatis* genomic DNA serially diluted 100 fold (Table 3A) and 10-fold (Table 3B) and used as template for the *C. trachomatis*-IMRS and 16S RNA PCR. Fig 3A shows the probit plot for the *C. trachomatis*-IMRS PCR assay, and Fig 3B shows the probit plot for the gold standard 16S rRNA PCR assay. The LLOD was calculated as the concentration at which *C. trachomatis* DNA can be detected with 95% confidence. Probit analysis estimation for *C. trachomatis*-IMRS PCR, Coefficient χ = -3.6494, *P*-value 0.7043 (Table 3C). As indicated, the IMRS primers for *C. trachomatis* had an LLOD = 9.5 fg/μL, Fig 3A. Probit analysis estimation for 16S rRNA PCR χ = -7.2101, *P*-value 0.9978, Table 3C. As indicated, gold standard primers for *C. trachomatis* had an LLOD = 4.31 pg/μL. Table 3C shows the statistics obtained from the Probit analysis indicating that the *C. trachomatis*-IMRS PCR assay had increased sensitivity compared to the gold standard 16S rRNA PCR assay.

**Table 3:** *Chlamydia trachomatis* genomic DNA was serially diluted 100 folds (A) and 10-folds (B) and used as template for the *C. trachomatis*-IMRS and 16S rRNA PCR to estimate the lower limit of detection. Table (C) shows the statistics obtained from the Probit analysis.

**A**
| Serial dilutions (pg/μl) | Replicates (5) |
| --- | --- |
| 100 | 5/5 |
| 1 | 5/5 |
| 0.01 | 4/5 |
| 0.0001 | 5/5 |
| 0.000001 | 0/5 |
| 0.00000001 | 0/5 |

**B**
| Serial dilutions (pg/μl) | Replicates (5) |
| --- | --- |
| 100 | 5/5 |
| 10 | 4/5 |
| 1 | 3/5 |
| 0.1 | 1/5 |
| 0.01 | 1/5 |
| 0.001 | 0/5 |

**C**
| Assay | $\chi$ Coefficient | P-Value |
| --- | --- | --- |
| 16S rRNA | -7.2101 | 0.9978 |
| IMRS | -3.6494 | 0.7043 |

**Fig 3:**
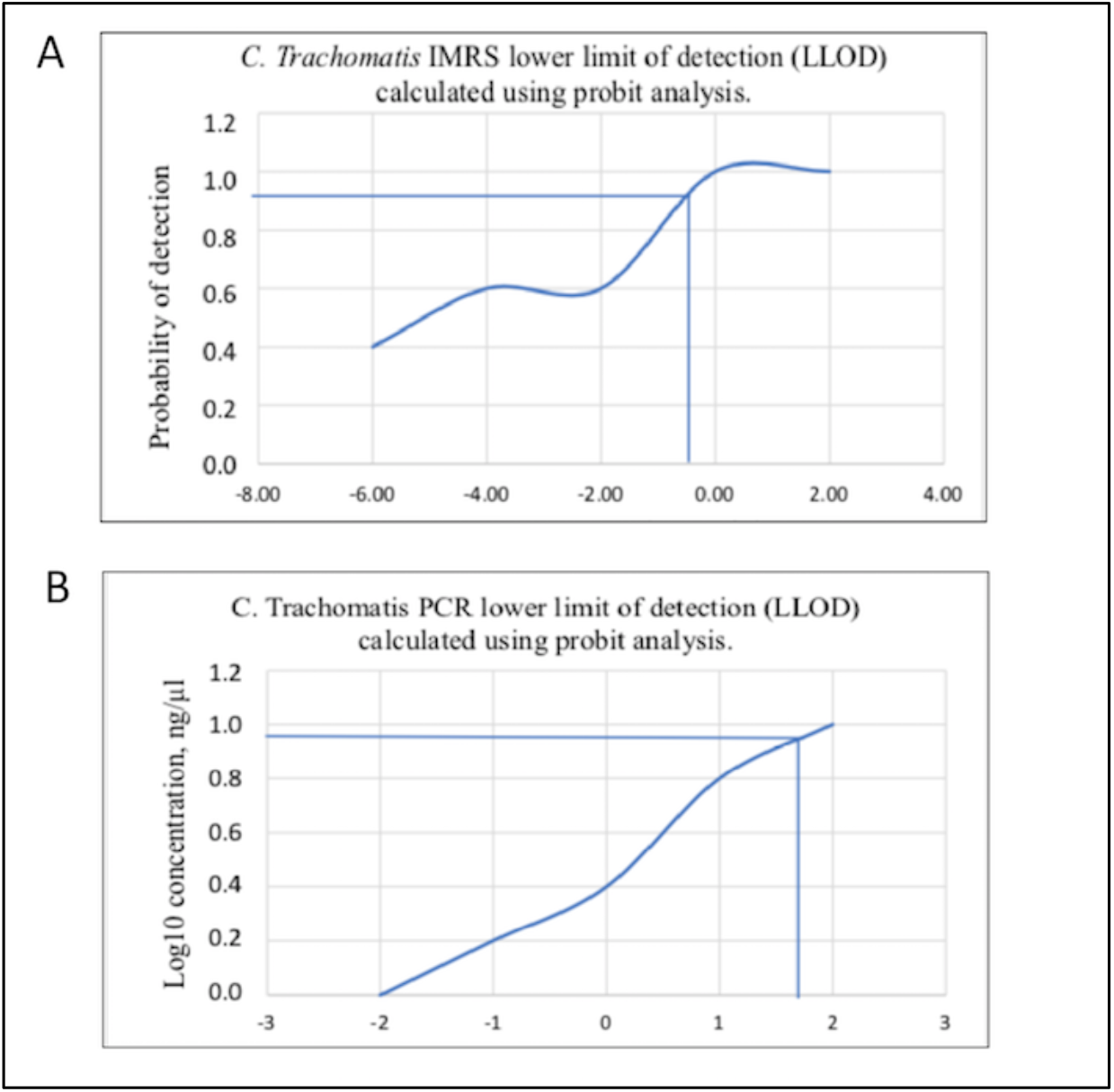
Probit regression analysis to estimate the lower limit of detection for the *C. trachomatis*-IMRS primers and the 16S rRNA PCR. Probit analysis estimation for *C. trachomatis*-IMRS (A). As indicated, the IMRS primers for *C. trachomatis* had an LLOD = 9.5 fg/μl. B: Probit analysis estimation for 16S rRNA PCR. As indicated, gold standard primers for for *C. trachomatis* had an LLOD = 4.31 pg/μl.

### 4.4 Isothermal Amplification of Genomic *Chlamydia trachomatis* DNA

Serially diluted genomic DNA was used to perform Iso-*C. trachomatis*-IMRS isothermal amplification. As shown in Fig 4A, the reaction products were visualized on a 1% gel. The Iso-*C. trachomatis*-IMRS assay successfully amplified *C. trachomatis* DNA down to 1.64×102 genome copies/μL using isothermal amplification. The LLOD for the *C. trachomatis*-Iso-IMRS assay was estimated at 0.3162 ng/μL (Fig 4C).

**Fig 4:**
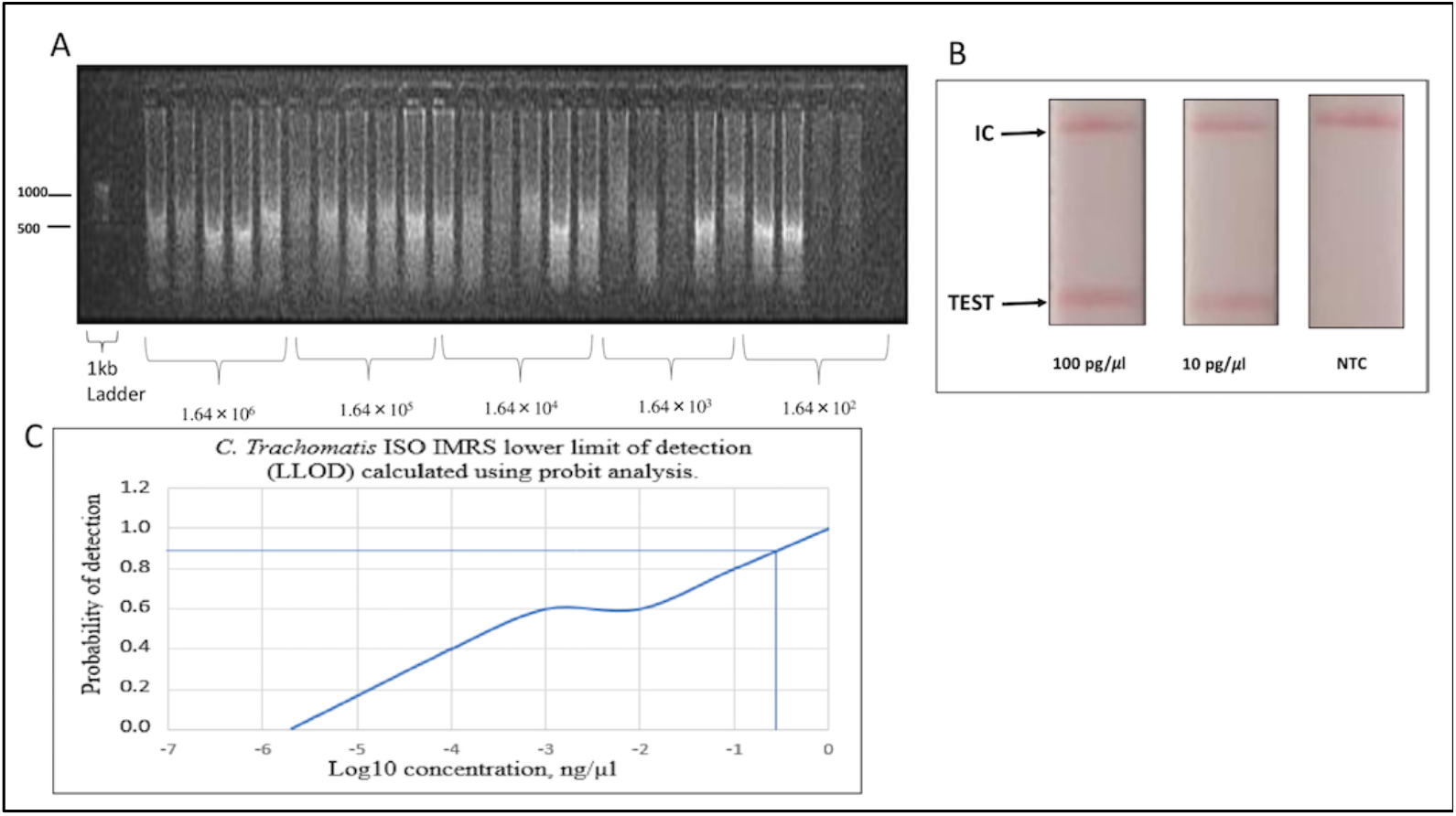
*C. trachomatis*-Iso-IMRS, lateral flow assay and estimation of the lower limit of detection of the isothermal assay. (A) Gel image of *C. trachomatis*-Iso IMRS assay products visualized on 1% gel. 5 replicates of each dilution served as DNA template for the *C. trachomatis*-Iso IMRS. DNA concentration is in genome copies per μl. (B) Visual read-out detection of serially diluted *Chlamydia trachomatis* DNA using the lateral flow assay. Amplicons were incubated at 65ºC for 1 hour and transferred onto strips as indicated. IC – Internal Control, NTC – Non Template Control. (C) Probit analysis estimation for *C. trachomatis* Iso-IMRS. As indicated, the IMRS primers for *C. trachomatis* had an LL0D = 0.3162 ng/μl.

### 4.5 Chlamydia trachomatis Lateral Flow Assay (LFA)

A visual readout of diluted genomic DNA was developed using modified primers and labelled probes, as shown in Fig 4B. The LFA readout of the amplification products was successful indicating the potential for the Iso-IMRS-based assay to be used in field settings.

### 4.6 Plasmid *C. trachomatis*-DNA concentration in ng/μl of transformed *E. coli* cells

To validate the exact regions that were amplified by the *C. trachomatis*-IMRS primers, we cloned the amplicon into blunt cloning vectors and transformed into electrocomperent *E. coli* cells and plated onto agar plates. DNA from eight (Figure 5) transformed *E. coli* cells was extracted and sequenced. Multiple sequencing alignment confirmed *C. trachomatis* sequences. These results suggested that the *C. trachomatis*-IMRS primers were specific for targets within the genome.

**Fig 5:**
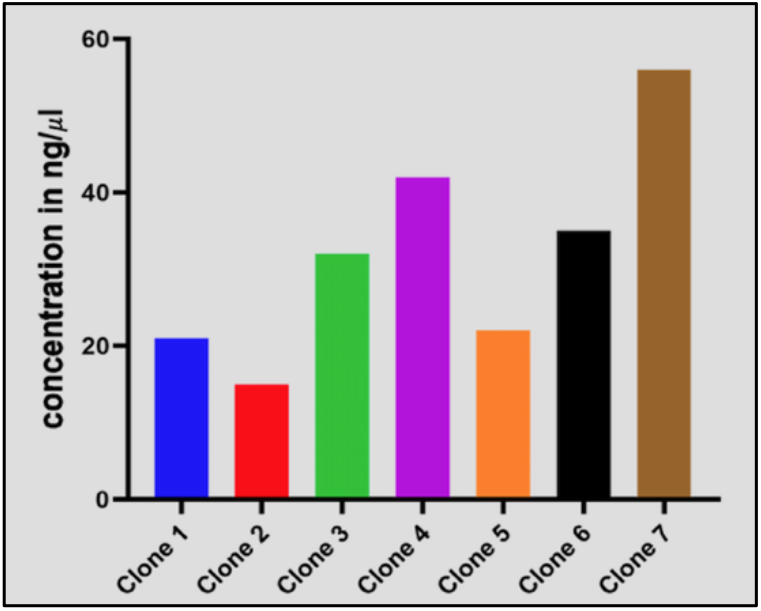
Transformed *E.coli* cells expressing *Chlamydia trachomatis* sequences amplified using *C. trachomatis*-IMRS primers.

### 4.7 Validation of the *C. trachomatis*-IMRS primers using clinical samples

A total of 16 *Chlamydia trachomatis* positive DNA samples from a cross-sectional study at the Kenyatta National Hospital were used to evaluate the reliability of the *C. trachomatis*-IMRS primers for detecting *C. trachomatis* DNA using RT-PCR assay as shown in Fig 6. The demographic information for recruited participants has been described in Table 4. The *C. trachomatis*-IMRS primers showed excellent concordance with the results obtained from conventional PCR diagnosis, successfully identifying *C. trachomatis* infections in infected samples.

**Table 4:** Demographic data for participants.

| Variable | Chlamydia positive,<br>n = 16,(%) | Chlamydia negative,<br>n = 187, (%) | P-Value |
| --- | --- | --- | --- |
| Age Category |  |  |  |
| 10 - 19 | 0, 0 | 2, 1 | 0.9467 |
| 20 - 29 | 6, 38 | 63, 34 |  |
| 30 - 39 | 9, 56 | 105, 56 |  |
| 40 - 49 | 1, 6 | 17, 9 |  |
| Marital status |  |  |  |
| Married | 13, 81 | 161, 86 | 0.5949 |
| Not Married | 3, 19 | 26, 14 |  |
| Level of Education |  |  |  |
| Primary | 1, 6 | 14, 7 | 0.5013 |
| Secondary | 8, 50 | 66, 35 |  |
| Tertiary | 7, 44 | 107, 57 |  |
| Employment status |  |  |  |
| Employed | 8, 50 | 55, 29 | 0.0875 |
| Not employed | 8, 50 | 132, 71 |  |
| Parity |  |  |  |
| Primipara | 1, 6 | 39, 21 | 0.3702 |
| Multipara | 14, 88 | 138, 74 |  |
| Grandmultipara | 1, 6 | 10, 5 |  |
| Gestational Age |  |  |  |
| First trimester | 0, 0 | 11, 6 | 0.0637 |
| Second trimester | 7, 44 | 37, 20 |  |
| Third trimester | 9, 56 | 139, 74 |  |
| HIV Status |  |  |  |
| Positive | 1, 6 | 1, 1 | 0.0263 |
| Negative | 15, 94 | 186, 99 |  |
| Miscarriage |  |  |  |
| None | 9, 56 | 111, 59 | 0.8421 |
| Once | 5 | 52, 28 |  |
| Twice | 2 | 14, 7 |  |
| Thrice | 0, 0 | 7, 11 |  |
| Quadruple | 0, 0 | 3, 2 |  |

**Fig 6:**
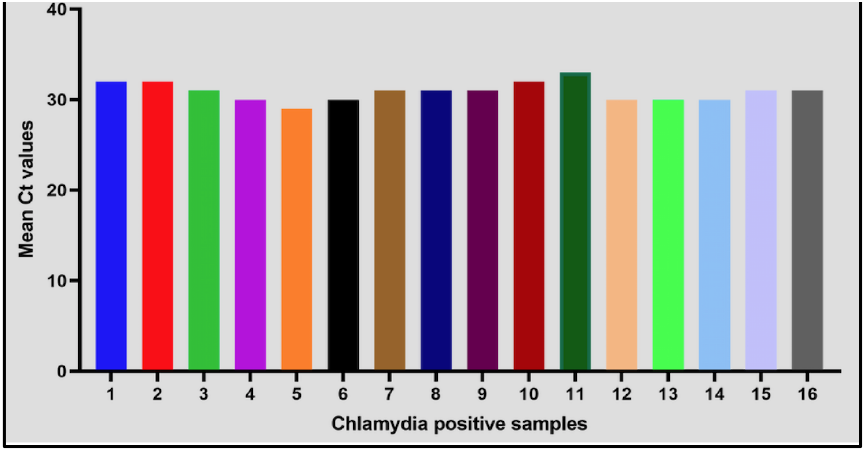
Mean Ct values of PCR confirmed 16 clinical DNA samples from RT-PCR assay that were used to validate the *C. trachomatis*-IMRS PCR primers.

### 4.8 Specificity and sensitivity of the *C. trachomatis*-IMRS primers

As shown in Fig 7, the *C. trachomatis*-IMRS primers were specific only for C. trachomatis DNA after amplification of *Treponema pallidum* (TP) and *Trichomonas vaginalis* (TV) genomic DNA using *C. trachomatis*-IMRS primers. *C. trachomatis* genomic DNA was used as a positive control. Also, compared to the gold standard conventional 16S-rRNA PCR, the *C. trachomatis*-IMRS PCR reliably detected genomic DNA at a concentration of 1 fg/μl (Fig 1B).

**Fig 7:**
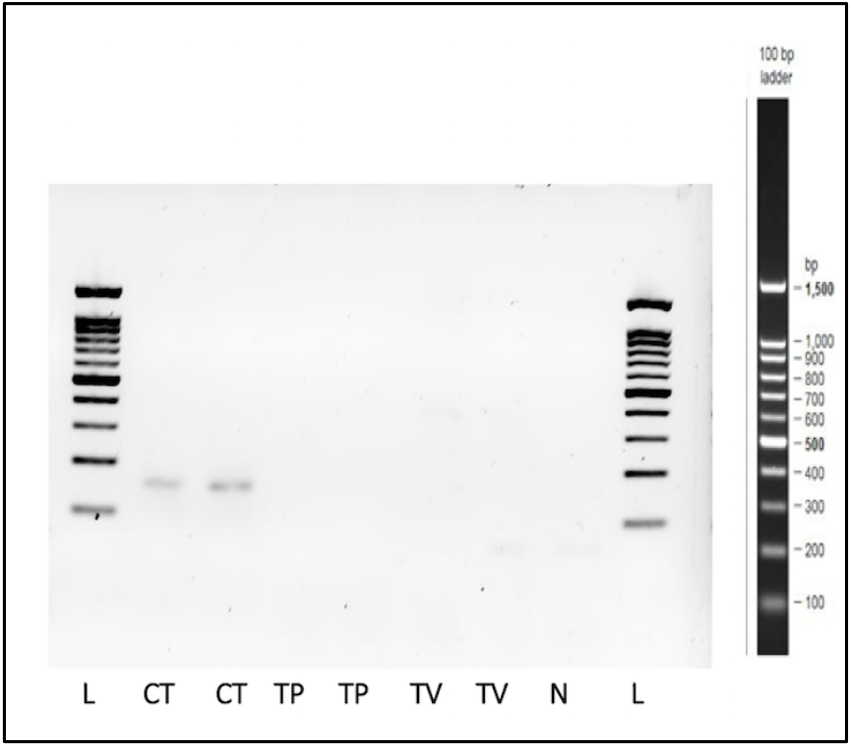
Gel image of PCR products obtained after amplification of *Treponema palidum* (TP) and *Trichomonas vaginalis* (TV) genomic DNA using *C. trachomatis*-IMRS primers. *C. trachomatis* genomic DNA was used as a positive control. N = Negative control, L = Ladder.

## 5. Discussion

Traditional techniques for identifying *C. trachomatis* infections have low sensitivity, require the collection of invasive specimens, require long duration for performance and reporting, and are expensive (17). As a consequence, there is a need to develop novel tests that are sensitive and specific. In the present study, we demonstrate the use of a deep genome mining strategy to identify identical multiple repeat sequences that could be used as robust primers for a novel nucleic acid-based test that is a highly sensitive test against *C. trachomatis*. Specifically, IMRS forward and reverse primers initiate amplification at multiple loci across the genome. This, in turn, improves the overall analytical sensitivity by generating more amplicons (21,22). We observed that, compared to the gold standard 16S rRNA PCR, amplification of specific sequences on the *C. trachomatis* genome using IMRS primers was more sensitive, generating a large number of amplicons of varying sizes hence a lower limit of detection of 0.0095 pg/mL (8.4 genome copies/mL). Our results are comparable to the Chlamydial Roche Amplicor Real-Time Quantitative PCR with a detection limit of 200 genome copies/ml (23). However, this assay targets up to 10 copies of chlamydial plasmid.

We further showed that isothermal amplification using IMRS PCR primers is reliable and sensitive for detecting *C. trachomatis*. The *C. trachomatis*-Iso-IMRS assay detected DNA up to 8.9 genome copies per mL. The Iso-IMRS assay was more sensitive than a Loop-mediated isothermal assay based on the ompA and orf1 genes which reported a limit of detection of 50 copies per mL (24).

Our study successfully developed and tested a novel Lateral Flow Assay testing platform to identify *C. trachomatis* to a concentration of 10 pg/mL (8.8 genome copies/ml) (Fig 6). Our finding was different from a study that developed a visual read-out of *C. trachomatis*-LAMP results, a gold nanoparticle-based lateral flow biosensor that reported a detection limit of 50 copies/ml after an incubation of 45 minutes (20).

Compared to nucleic acid methods that are used for the detection of *C. trachomatis* that are mostly suboptimal (22), the *C. trachomatis*-IMRS PCR primers were specific and sensitive when used for the identification of *C. trachomatis* DNA.

## 6. Conclusion

Put together; here we show that the IMRS algorithm can serve as a platform technology for designing primers that are sensitive and specific for *C. trachomatis*. This platform has potential application in other bacterial and non-bacterial pathogens and could significantly improve future disease diagnostics procedures.

## Data Availability

All data produced in the present work are contained in the manuscript

https://www.dropbox.com/s/2lzkgmx10tsfnz5/NG_Data_F.pptx?dl=0

## Ethical Statement

Use of these samples and study proposal was approved by the Mount Kenya University Ethical Review Committee (MKU/ERC/1649).

## Data Accessibility

All data generated or analysed during this study are included in this publishedarticle [a supplementary information files].

DNA sequences: Genbank accessions AE001273 AE001275 AE https://www.ncbi.nlm.nih.gov/nuccore/AE001273.1/

## Authors’ Contributions

C.S.: conceptualization, data curation, formal analysis, methodology, writing—original draft; S.K.: data curation, formal analysis, investigation, methodology, supervision, visualization, writing— original draft, writing—review and editing; B.N.K.: investigation, writing—review and editing; R.K.: resources, writing—review and editing; M.M.: resources, writing—review and editing; H.W.: resources, writing—review and editing; M.K.: resources, writing—review and editing; I.N.: resources, writing—review and editing; H.M.A.: resources, writing—review and editing; B.O.: funding acquisition, writing—review and editing; N.P.: resources, writing—review and editing;. J.D.: writing— review and editing; C.M.K.: resources, writing—review and editing; S.R.L.: methodology, writing— review and editing; J.G.: conceptualization, funding acquisition, investigation, methodology, project administration, resources, supervision, writing—original draft, writing—review and editing.

## Acknowledgement

We thank Dr. Karen Muthembwa and all clinicians working at the STI Clinic in Kenyatta National Hospital

